# Effectiveness of maternal vaccines and long-acting monoclonal antibodies against respiratory syncytial virus hospitalisations in early life: a scoping review of dynamic modelling studies

**DOI:** 10.1101/2025.04.16.25325979

**Authors:** Alessandra Bicego, James G Wood, Anthony T Newall, Alexandra B Hogan

## Abstract

**Background:** Respiratory syncytial virus (RSV) is a leading cause of respiratory illness and hospitalisation in infants and young children. New pharmaceutical interventions for preventing severe RSV in early life, namely a maternal vaccine and a long-acting monoclonal antibody, have recently been approved and are now available for use. Over the past decade, mathematical models of RSV transmission have been used to predict the impact of novel pharmaceutical interventions, in anticipation of future product licensure, and to model the potential impact of newly available interventions. However, these models have varied in structure, parameterisation, assumptions, and the immunisation schedules simulated.

**Methods:** In this scoping review, we surveyed published dynamic modelling studies that estimated the prospective population-level impact of either an RSV maternal vaccine or a long-acting monoclonal antibody in children <2 years, focussing on upper-middle- and high-income settings. We extracted data on the model structures, assumptions, and parameterisation, and synthesised the modelled estimates of future immunisation impact across studies.

**Findings:** Of the 210 articles reviewed, a total of 7 studies met our criteria. Two studies modelled only a maternal vaccination strategy, one modelled a long-acting monoclonal strategy, and four modelled both. Estimates ranged from 5–21 and 11–32 annual RSV hospitalisations per 1,000 children averted for a maternal vaccine and a monoclonal antibody respectively in infants aged <3 months, corresponding to ranges of approximately 10–53% and 32–70% hospitalisations averted. Six of the studies explicitly captured natural maternally-derived immunity in infants following birth, but the magnitude and duration varied widely.

**Interpretation:** All studies found that either a maternal vaccine and/or a long-acting monoclonal antibody could significantly reduce RSV hospitalisations in children younger than 12 months. We identified broad consistency in results across studies, and all studies captured declining impact in older children. Predicted impact was larger for a monoclonal antibody compared to a vaccine, due to higher assumed coverage and efficacy. Given assumptions around maternal immunity varied widely, improving both models and the evidence base for this process would be beneficial.

## Introduction

Respiratory syncytial virus (RSV) is a major cause of respiratory morbidity in young children and infants and presents a substantial global health burden (1,2). Infection is common (with almost all children thought to be infected by the age of two years) and recurs throughout life. The highest incidence of severe RSV disease occurs in young children. Close to 40% of hospitalisations due to severe RSV disease occur within the first six months of life, and RSV is associated with a larger hospitalisation burden than influenza in children younger tha5 years (2–6).

Several new products, designed to prevent severe RSV in infants, have recently progressed through the clinical trials pipeline. Abrysvo, a maternal vaccine that can be delivered in the final trimester of pregnancy, was approved by the US Food and Drug Administration (FDA) for use in pregnant women in August 2023 (7). The phase 3 clinical trial for Abrysvo indicated efficacy against medically-attended RSV-associated lower respiratory tract illness (MA-RSV-LRTI) over 90 days of 57.1% (99.5% confidence interval 14.7 to 79.8) (8). Nirsevimab (Beyfortus) is a long-acting monoclonal antibody therapy, to be delivered to infants or young children either in the first few days of life (ideally before hospital discharge) or in advance of an RSV season. Nirsevimab was approved by the FDA July 2023 (9). Clinical trial data for nirsevimab indicated efficacy against MA-RSV-LRTI in the first 150 days after injection of 79.5% (95% CI 65.9 to 87.7) (10). Vaccines are now available for prevention of RSV disease in older adults (11), and paediatric vaccines for administration to young children are in development (12).

While clinical trials provide direct efficacy results for symptomatic RSV and more severe outcomes, the extent to which vaccines and monoclonals prevent acquisition of RSV infection, and the longer-term durability of protection, are less well known. In addition, there remain many unknowns about natural RSV immunity in early-life; while RSV-specific protective antibodies are known to transfer transplacentally in utero, a full quantitative understanding of how these antibodies translate to early life protection is not yet available (13). Mathematical models are frequently used to simulate and predict disease dynamics in the context of novel vaccines and therapeutics, including before empirical studies such as clinical trials have been completed, while exploring a range of hypothetical uptake, efficacy, or scheduling scenarios (14). However, different modelling studies will vary in approaches to model structure, parameterisation, and design of scenarios. These differing assumptions can strongly influence both the simulated epidemiological impact and cost-effectiveness of potential interventions, with the potential to cause confusion for decision-makers.

In the context of RSV prevention in infants, we are currently in a unique position, with two competing products available with different characteristics and indications, and a need for health decision-makers to recommend the most appropriate and effective programs for RSV prevention in their policy setting. Mathematical models can form part of this decision-making process. The purpose of our study was to perform a scoping review of the published literature on prospective population-level dynamic mathematical modelling studies that estimated the impact of novel pharmaceutical interventions on RSV disease in infants and young children. Using the reviewed studies, we aimed to collate information on the range of model structures and assumptions, with a particular focus on vaccine and monoclonal characteristics and modelled scenarios, and on assumptions about natural immunity in early life. By synthesising modelled outputs across studies, we aimed to capture the variation in the predicted impacts of a maternal vaccine and monoclonal antibody in the first two years of life.

## Methods

### Search strategy and screening

For our scoping review, on 11/07/2024 we conducted a systematic search of EMBASE for peer-reviewed original research articles published between 01/07/2014 and 30/06/2024. We aimed to capture articles that focused on the application of compartmental differential equation models (of the Susceptible-Exposed-Recovered-Susceptible (SIRS) form) to simulation of RSV transmission in humans and related health/economic outcomes, under scenarios involving either a maternal vaccine, or a long-acting monoclonal therapy. The list of search terms is in the Supplementary material. Because RSV hospitalisation rates, as well as contact patterns and demographics, can be quite different between higher and lower resourced settings, we focused only on studies pertaining to upper-middle or high-income settings (based on The World Bank income classification). We excluded any studies that focused specifically on the impact of the COVID-19 pandemic on RSV epidemiology. Unpublished studies, review papers, small case studies, non-English language studies and conference abstracts were also excluded.

All screening was undertaken by one reviewer, with discussion with two other reviewers to resolve any uncertainty as to a study’s eligibility. We did not conduct a quality appraisal or risk of bias assessment for the included studies, as these are not requirements of a scoping review (15,16).

### Data extraction

From the final set of articles we extracted these data: manuscript metadata, study population, model age stratification, calibration data, research question, model type and structure, interventions modelled and assumptions about these interventions (schedule, mode of action, duration of protection, efficacy, and coverage), parameterisation of natural maternally-derived immunity in infants, outcome measures, headline results, and modelled impact stratified by age group. Results pertaining to additional product types (such as paediatric or elderly RSV vaccines) or relating to strategies focussed only on high-risk infant groups were not extracted.

### Data analysis

We summarised the extracted data by presenting the design and results in the context of three areas: (1) model structure, assumptions and parameterisation, (2) scenarios and projected impact of maternal vaccination and/or monoclonal therapy in children younger than two years, and (3) modelled mechanisms and assumptions regarding natural maternally-derived infant immunity in the model. We extract estimates of modelled impact in terms of either hospitalisations, incidence of severe RSV disease, or incidence of MA-RSV-LRTI (or similar). Extracted results were transformed to one or both of the following metrics: averted RSV hospitalisations per 1,000 children by age, and averted RSV hospitalisations as a percentage reduction in that age group. Where results were only available in a figure the data were extracted using an online plot digitiser (17).

#### Availability of data

The extracted impact data and code to reproduce the results are available at https://github.com/abhogan/rsv_model_scoping.

## Results

A total of 210 articles were identified, of which 22 studies were excluded during pre-screening (duplicates, reviews, or not peer-reviewed), a further 131 were excluded following title screening, and 28 and 22 articles excluded respectively at the abstract and full-text screening stages respectively (Figure 1). The most common reasons for exclusion included being use of a non-dynamic model, or analysis not being for an upper-middle or high-income setting. Seven articles remained for inclusion in the review (Tables 2 and 3). These included two studies by Hodgson and colleagues (18,19), featuring the same underlying transmission model; we included both studies as the model parameterisation and scenario design differed substantially between the studies, but excluded a third study from the same group (Hodgson 2022 (20)) due to limited differences in parameterisation and scenario design compared to the 2024 Hodgson paper (19). The papers by Hogan et al and Prasad et al also used a similar underlying model structure, but had substantial differences in parameterisation and scenarios, and therefore both were included (21,22).

**Figure 1:**
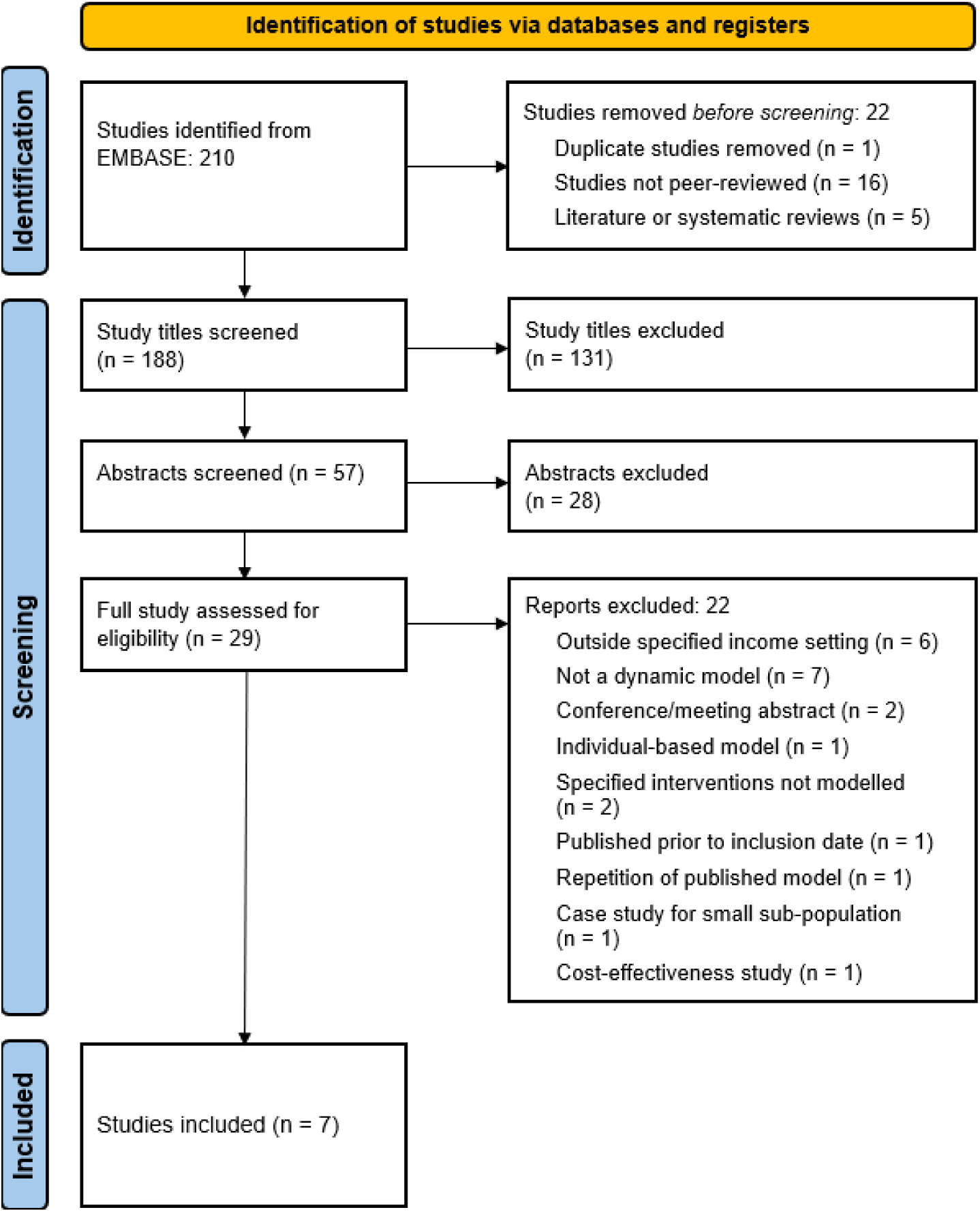
Preferred Reporting Items for Systematic Reviews and Meta-Analyses (PRISMA) flow diagram for inclusion of studies (41).

The studies included in the review generally aimed to model the impact of a novel maternal vaccine or long-acting monoclonal antibody on RSV disease in young children younger than two years of age, with a focus on the first six months of life (Tables S1 and S2). Settings included England and Wales, Australia, New Zealand, and the USA, published during 2017– 2024. Of the studies included for analysis, two modelled only a maternal vaccination strategy, one modelled a long-acting monoclonal strategy, and four studies modelled both (Tables 1 and 2). In these four studies (18,19,22,23), strategies used monoclonal or maternal vaccines in isolation, and did not consider the impact of a combined immunisation program.

**Table 1:** Maternal vaccination studies. Summary of model structures, assumptions and parameters for the reviewed studies that modelled the impact of maternal vaccination programs. In the transmission model structure column, compartments are represented by S (susceptible), E (exposed), I (infectious, or symptomatic infection in Hodgson), R (recovered), A (asymptomatic infection), and M (maternally-derived protection). The transmission model structure does not describe the additional observation model states for detected, hospitalised, or more severe classes that were included in all models. All models included in the review were stratified by age. Parameters relating to product and delivery assumptions outlined in the table relate to the main (or default) set of parameters in each study; additional sensitivity analyses may have been performed but assumptions and outputs were not extracted for these analyses. Note that the model structures in Hodgson 2020 and Hodgson 2024 are largely the same, but parameters and delivery assumptions varied between the studies. In addition, the models in Prasad 2021 and Hogan 2017 are similar (with overlap in authors), but again parameters and scenarios were different.

| Study | Country | Transmission model structure | Product assumptions |  |  | Delivery assumptions |  |
| --- | --- | --- | --- | --- | --- | --- | --- |
|  |  |  | Mode of action | Efficacy | Average duration of protection (days) | Schedule | Coverage |
| Hodgson 2020 (18) | England and Wales | MSEIARS with infection history | Efficacy against symptomatic infection; additional efficacy against hospitalisation* | Infection: 41.4%<br>Hospitalisation: 53.5% | 133.5 | Seasonal; year-round | 60% |
| Hodgson 2024 (19) | England and Wales | MSEIARS with infection history | Efficacy against infection; additional efficacy against hospitalisation* | Infection: 49.8% (8) | 180 | Seasonal; year-round | 60% |
| Hogan 2017 (21) | Australia | SEIRS; maternally derived protection in infant S classes | Efficacy against infection | 80% (25) | 180 | Year-round | 50% |
| Prasad 2021 (22) | New Zealand | SEIRS; maternally derived protection in infant S classes | Efficacy against infection | 40% first 3 months, 20% thereafter (27) | 180 | Year-round | 50% |
| van Boven 2020 (24) | USA | SIR; discrete seasonal immunity process | Efficacy against infection | Combined with coverage | 180 | Year-round | 50% |
| Zheng 2022 (23) | USA | MSIS with infection history | Efficacy against infection | 85% (25,26) | 150 following waning of maternally induced protection | Seasonal; year-round | 85–95% |

\* Note that in Hodgson et al 2020 and Hodgson et al 2024, the models captured vaccine-derived protection to pregnant women themselves. Additionally, vaccinated mother-infant pairs were captured in the model compartment stratification, to reflect a reduced force of infection from vaccinated mothers upon their infants.

**Table 2:** Long-acting monoclonal antibody studies. Summary of model structures, assumptions and parameters for the reviewed studies that modelled the impact of long-acting monoclonal antibody programs. In the transmission model structure column, compartments are represented by S (susceptible), E (exposed), I (infectious, or symptomatic infection in Hodgson), R (recovered), A (asymptomatic infection), and M (maternally-derived protection). The transmission model structure does not describe the additional observation model states for detected, hospitalised, or more severe classes that were included in all models. All models included in the review were stratified by age. Parameters relating to product and delivery assumptions outlined in the table relate to the main (or default) set of parameters in each study; additional sensitivity analyses may have been performed but assumptions and outputs were not extracted for these analyses. Note that the model structures in Hodgson 2020 and Hodgson 2024 are largely the same, but parameters and modelled scenarios varied between the studies.

| Study | Country | Transmission model structure | Product assumptions |  |  | Delivery assumptions |  |
| --- | --- | --- | --- | --- | --- | --- | --- |
|  |  |  | Mode of action | Efficacy | Average duration of protection (days) | Schedule | Coverage |
| Hodgson 2020 (18) | England and Wales | MSEIARS with infection history | Efficacy against symptomatic infection; additional efficacy against hospitalisation | 70.1% symptomatic infection; 78.4% hospitalisation | 275 | Seasonal: Infants born during season Oct-Feb<br>Seasonal +: Infants born Oct-Feb, plus all infants <6 months during Oct | 90% |
| Hodgson 2024 (19) | England and Wales | MSEIARS with infection history | Efficacy against infection | 77.3% (10) | 150 | Seasonal: Infants born Sept-Feb<br>Seasonal +: Infants born Sept-Feb, plus all infants 1-6 months during Sept<br>Year-round at birth | 90% |
| Prasad 2021 (22) | Australia | SEIRS; maternally derived protection in infant S classes | Efficacy against infection | 50% (28) | 150 | Seasonal: Infants aged <6 months entering RSV season or born during season | 50% |
| Voirin 2022 (29) | USA | MSIRS with infection history | Efficacy against severe RSV-LRTI (scenario 1); additional efficacy against transmission (viral shedding) (scenario 2) | 70% (scenario 1); additional 50% against transmission (scenario 2) (28) | 150 | Seasonal: Infants <7m entering RSV season or born during season | 71% |
| Zheng 2022 (23) | USA | MSIS with infection history | Efficacy against infection | 100% (28) | 275 | Seasonal: Infants <6 months entering RSV season or born during season | 85–95% |

### Model structures and outputs

All studies presented age-stratified model structures of the broad Susceptible, Exposed, Infectious, Recovered, Susceptible (SEIRS) form. The two Hodgson et al studies additionally stratified the infectious class by symptomatic status (18,19), and four of the seven studies further stratified susceptibility via additional compartments to capture the reduced risk of infection and/or transmission after second and subsequent infections (Tables 1 and 2).

Studies generally reported outputs per age group as the annual number of averted RSV hospitalisations per multiple of 1,000 children and/or the percentage reduction in RSV hospitalisations. The study by van Boven et al reported the main outputs in terms of impact on the annual infection attack rate in individuals aged 0, 1–4, 5–9 and 65+ years, rather than an impact upon severe disease (24).

### Model assumptions, parameterisation and scenarios

Transmission models were calibrated using time series data of age-specified hospitalised or medically-attended laboratory-confirmed RSV cases in young children, with age-specific reporting rates (i.e. the fraction of RSV infections that are tested and hospitalised) estimated as part of the model calibration process (Table S4).

Assumptions relating to vaccine characteristics in earlier studies were generally informed by the published WHO Preferred Product Characteristics for RSV Vaccines (21,23,25), and later studies were informed by clinical trial data (8,26,27). We found variation in the translation of trial results to model efficacy parameters (including the magnitude of efficacy against infection versus hospitalisation, and the duration of protection), with average efficacy against infection varying from approximately 40–85% (Table 1). For models of the a long-acting monoclonal antibody, most were informed by nirsevimab clinical trial results (28), and average efficacy against infection varied from 50–100% (Table 2). Duration of protection was relatively consistent across the studies, with an average of 4–6 months for the maternal vaccine (Table 1), and 5–9 months for the monoclonal (Table 2). Note that in Hodgson et al 2024, efficacy was implemented as a time-varying probability of protection, based on fitting to data from clinical trial studies (19). Additionally in the Hodgson et al 2020 and Hodgson et al 2024 studies, the models captured vaccine-derived protection to pregnant women themselves. Further, vaccinated mother-infant pairs were captured in the model compartment stratification and contact structure, to represent the reduced force of infection from vaccinated mothers upon their infants (18,19).

Immunisation scheduling and coverage assumptions varied widely between the studies. All maternal vaccination studies modelled year-round programs, and the two Hodgson et al studies and Zheng et al additionally simulated seasonal programs (18,19,23). For the long-acting monoclonal antibody, studies generally assumed a seasonal program where infants born during a defined “RSV season” were immunised, and older infants up to 6 months of age immunised in advance of the season (18,19,22,29). Base-case immunisation uptake varied between 50– 95%, with a range of coverage values generally tested within each study (Tables 1 and 2).

### Modelled epidemiological impact

For a maternal vaccine, using the base-case results from each study only, the median hospitalisations or MA-LRTI averted across the studies was 8 per 1,000 children aged younger than 3 months (range 5–21), and 3 per 1,000 children aged between 3 to 5 months (range 1–12). This translated to median reductions of 20% (range 10–53%) and 11% (range 3–40%) in these groups respectively. Note that the level of coverage varied between studies, and while 6 of the 7 studies assumed coverage values of 50% or 60%, the Zheng et al study used coverage values of 85–95%, and reported the highest magnitude of impact (see Figure 2). The study by van Boven et al 2020 reported impact as change in the infection attack rate, estimating a 27% reduction in 0–1-year-old children and a 10% increase in 1–4-year-old children.

**Figure 2.**
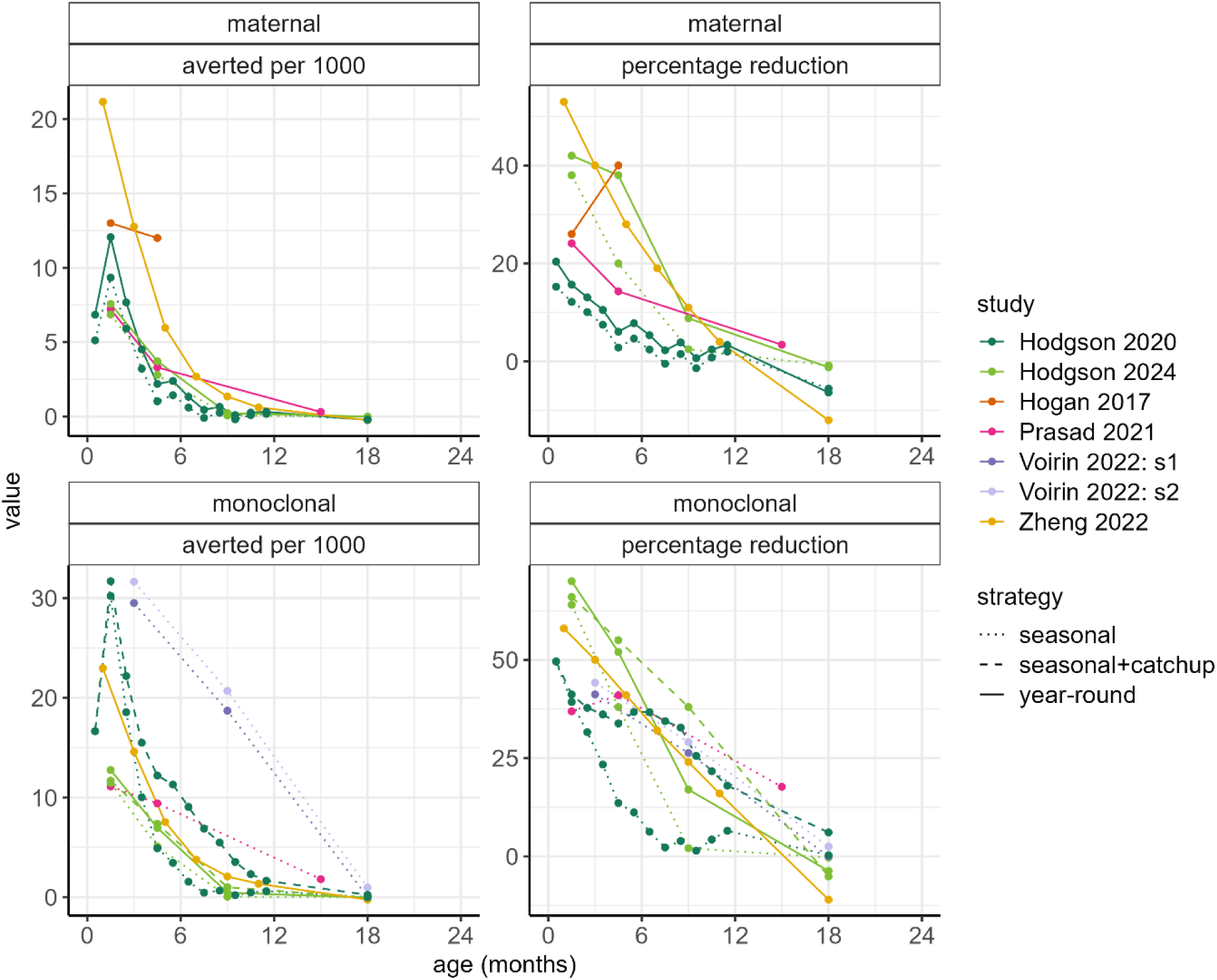
Modelled estimates of maternal vaccine impact (upper row) and long-acting monoclonal antibody impact (lower row) in children up to two years of age, by age group in months. In each panel, the left-hand side figure shows the modelled number of averted hospitalisations per 1,000 children, and right-hand side figure shows the percentage reduction in RSV hospitalisations. Studies are denoted in coloured points and lines, where each point estimate represents the value at the midpoint of the age group reported. Results were extracted from reviewed studies (either from tables, figures, or raw results files where provided), where the results were presented in an age-stratified metric that allowed this comparison and were transformed to the relevant scale where required. For the Voirin et al study (29), impact is shown in terms of RSV medically-attended lower respiratory tract infections, rather than RSV hospitalisations. The two sets of modelled outputs for the Voirin et al study (29) relate to no assumed efficacy against viral shedding (scenario 1: s1) and additional efficacy against viral shedding (scenario 2: s2). Note that coverage and efficacy assumptions varied between these studies (See Tables 2 and 3 for additional information about model assumptions and data sources).

For a long-acting monoclonal antibody, across both year-round and seasonal implementation strategies and across studies, the median estimated numbers of averted hospitalisations or RSV-MA-LRTI per 1,000 children <3 months of age was 17 (range 11–32) per 1,000, corresponding to a percentage reduction of 50% (range 32–70%). For children aged 3–6 months, the median averted hospitalisations was 8 per 1,000 (range 3–15) corresponding to a percentage reduction of 37% (range 11–55%). respectively. For children 12–24 months of age (after the assumed duration of protection for both interventions had passed), models predicted either minimal changes, or a small increase in total RSV hospitalisations (due to a delay in a child’s first RSV infection) for both maternal vaccination and long-acting monoclonal antibody scenarios (Figure 2).

While in this scoping review we focussed on the impact of immunisation strategies in children in the first two years of life, we noted the extent to which models reported herd effects of RSV interventions. The studies by Hogan et al, Prasad et al, and Zheng et al did not report impact of interventions in older ages (21–23), and Van Boven et al reported a negligible impact in older adults following maternal vaccination (24). Hodgson et al 2024 reported a minimal impact across all modelled intervention scenarios in the population older than five years of age. Voirin et al, for the scenario in which the long-acting monoclonal was assumed to reduce viral shedding, reported a larger herd effect compared to other studies, and estimated an average reduction in RSV-MA-LRTIs of 6.9% in adults >65 years of age (29). In both Hodgson et al 2020 and Hodgson et al 2024, the model compartmental structure captured the protective effect of vaccination in pregnant women themselves (18,19).

### Capturing natural maternally-derived immunity

Of the reviewed studies, two assumed a period of temporary but complete natural maternally-derived immunity, with this period of protection ranging from an average of 2 to 4 months between studies (23,29) (Table S3). In the two Hodgson et al studies, immunity in infants was dynamically linked to the proportion of adult women of childbearing age, who have recently been infected (18,19). The studies by Hogan et al and Prasad et al assumed a period of temporary partial immunity, with susceptibility to infection reduced by a scaling parameter in the first three months of life, to capture temporary partial protection against infection and severe disease (21,22). The study by van Boven et al did not explicitly capture maternally-derived protection in infants (24).

## Discussion

In this scoping review we aimed to synthesise outputs from published dynamic compartmental transmission models to assess the future population-level impact of RSV immunisation strategies, focussing on high-income settings. We found that studies varied in terms of country setting, and assumptions relating to vaccine and monoclonal efficacies and durations. While compartmental structure and age-stratification were broadly consistent, only some studies stratified the population by history of infection, and the modelled temporary natural maternally-derived immunity in infants following birth varied between complete and partial. Despite these differences, all models predicted a substantial benefit from the introduction of a maternal vaccine or long-acting monoclonal antibody in the first six months of life, although the magnitude of impact varied between studies. Estimated impacts in children above one year of age were generally small.

The studies reviewed were prospective in nature, and generally published prior to approval and widespread rollout of novel RSV prevention products, although most studies were informed by efficacy data from phase 2 or 3 clinical trials. Even when clinical trial data were available, we found that efficacy data were incorporated into models in different ways; RSV clinical trials have reported efficacy against respiratory tract disease and more severe outcomes, therefore in models simulating transmission, additional assumptions are required in regards to protection against acquisition of infection versus development of disease. While we did not explicitly quantify the sensitivity of outputs to vaccine-specific parameters, it is worth noting that the efficacy and mode of action was only one source of variation between models.

An increasing number of countries have integrated RSV maternal vaccines and/or long-acting monoclonals into their immunisation programs, with program design being context-dependent and varying between jurisdictions. In the United States, the CDC recommends RSV maternal vaccination or monoclonal antibody administration (nirsevimab), during specific months only, to maximise protection during the RSV season (30) whereas in the United Kingdom, pregnant women are now offered the maternal RSV vaccine Abrysvo year-round, and nirsevimab offered to high-risk infants only (31). In Canada, a long-acting monoclonal antibody is preferred over the maternal RSV vaccine, and is recommended for any infant entering their first RSV season, and for infants at increased risk during their second RSV season (32). In Australia, Abrysvo and nirsevimab are now part of the National RSV Mother and Infant Protection Program (RSV-MIPP), with Abrysvo available on the National Immunisation Program for pregnant women at 28–34 weeks gestation, and nirsevimab available with administration varying by subnational health jurisdiction (33).

Studies are now beginning to emerge around population-level effectiveness of Abrysvo and nirsevimab in infants. Studies of the real-world benefit of nirsevimab have reported effectiveness against RSV hospitalisation in young children in the range of 70–90% (34–37), and population-level studies of Abrysvo benefit remain underway. While these studies capture differences in protection against severe disease between infants with and without receipt of passive RSV immunisation in the context of measured levels of uptake, they provide little information about protection against milder disease or infection itself. These latter outcomes are important in regard to any change more generally in risk of RSV infection, including under scenarios or coverage levels not currently used in practice. Here mathematical models retain an important role, with the emerging data potentially helping to refine predictions and guidance as to optimal RSV prevention strategies.

Previous reviews of mathematical models for RSV interventions exist (38–40), with the review by Treskova et al 2021 including a broader range of model types (not only dynamic transmission models) and finding an average estimated reduction in RSV hospitalisation of 27% from a maternal vaccine, and 50% for direct infant immunisation, which fall within the ranges reported in this study (40). The study by Mezei et al 2021 only considered lower-middle income countries, whereas we focussed on higher-income settings. The large review by Lang 2022 focussed on model design and parameterisation (38), whereas we applied a narrower set of criteria and aimed to synthesise the quantitative results across studies. As our scoping review was performed after the completion of phase 3 clinical trials of both maternal vaccine and long-acting monoclonal products, it therefore includes studies that account for these data.

This review highlighted several research gaps and opportunities for future investigation. First, the models in this review were generally calibrated to data on laboratory-confirmed hospitalisations, or primary care or emergency department presentations, with the age-specific proportion of infections resulting in hospitalisations inferred as part of the fitting routine. Globally, there are a lack of data on the age-specific prevalence and incidence of RSV infection in infants, and additional epidemiological evidence could better inform these estimates of age-stratified RSV transmission (40). Second, while some studies in this review modelled the independent impact of each of maternal vaccination and long-acting monoclonal antibody strategies, none of the identified studies modelled the total impact of a combined program. Modelling studies that estimate the total impact of planned immunisation programs (such as year-round maternal vaccination, supplemented by seasonal or high-risk monoclonal antibody administration), accounting for potential differential levels of community acceptance and uptake of each product, are needed to guide health policymakers on the anticipated effectiveness and efficiency of such programs. Finally, we found that the modelled assumptions about the magnitude and durability of maternally-derived immunity in infants varied widely between models, and that sensitivity of intervention impact to these assumptions was not consistently explored; this presents an avenue for future work.

There are several limitations to this review. The first is that reported outputs were not always consistent between studies, and that assumptions and parameters varied, making direct comparison difficult. Where possible, results were transformed to allow comparison, and differences between studies were acknowledged. Second, in this study we considered only upper-middle and high-income settings, to reduce heterogeneity in model assumptions, including demographic structures, contact patterns, burden and schedules modelled. From a model design and parameterisation perspective, models for lower-resourced settings may capture quite different contact patterns and demographic structures, and a disproportionately high burden of RSV is observed in young children in lower-resourced settings (2), therefore parameterisation and the scenarios modelled may vary from those shown here. Models of RSV transmission in lower-middle-income settings were the focus of a recent review by Mezei et al (39).

After several decades of research into RSV disease prevention, we now have the opportunity to make significant inroads into reducing respiratory morbidity and mortality in early life. Mathematical modelling studies such as those reviewed here can provide valuable evidence on the predicted impact of these interventions across a range of settings, and the ability to synthesise information on population-level impact generated by models will be beneficial for decisionmakers.

## Data Availability

The extracted impact data and code to reproduce the results are available at https://github.com/abhogan/rsv_model_scoping. All other data are available in the manuscript.

## Supplementary material

### Supplementary methods

Complete list of search terms used on EMBASE for article screening

1. Respiratory syncytial virus infection/ or respiratory syncytial virus pneumonia/
2. Human respiratory syncytial virus/
3. (rsv or hrsv or respiratory syncytial virus).mp. [mp=title, abstract, heading word, drug trade name, original title, device manufacturer, drug manufacturer, device trade time, keyword heading word, floating subheading word, candidate term word]
4. 1 or 2 or 3
5. Mathematical model/
6. Susceptible infected recovered model/
7. Susceptible exposed infectious recovered model/
8. Compartment model/
9. Stochastic model/
10. Epidemiological model/
11. (Mathematical model or dynamic model or mathematical modelling or SIR model or SEIR model or SEIRS model or transmission model or compartmental model or deterministic model or individual-based model or stochastic model or epidemiological model).mp. [mp=title, abstract, heading word, drug trade name, original title, device manufacturer, drug manufacturer, device trade time, keyword heading word, floating subheading word, candidate term word]
12. (model* adj4 (mathematical or dynamic or SIR or SEIR or SEIRS or transmission or compartmental or deterministic or individual-based or stochastic or epidemiological.mp. [mp=title, abstract, heading word, drug trade name, original title, device manufacturer, drug manufacturer, device trade time, keyword heading word, floating subheading word, candidate term word]
13. 5 or 6 or 7 or 8 or 9 or 10 or 11 or 12
14. 4 and 13

### Supplementary results

**Table S1:**
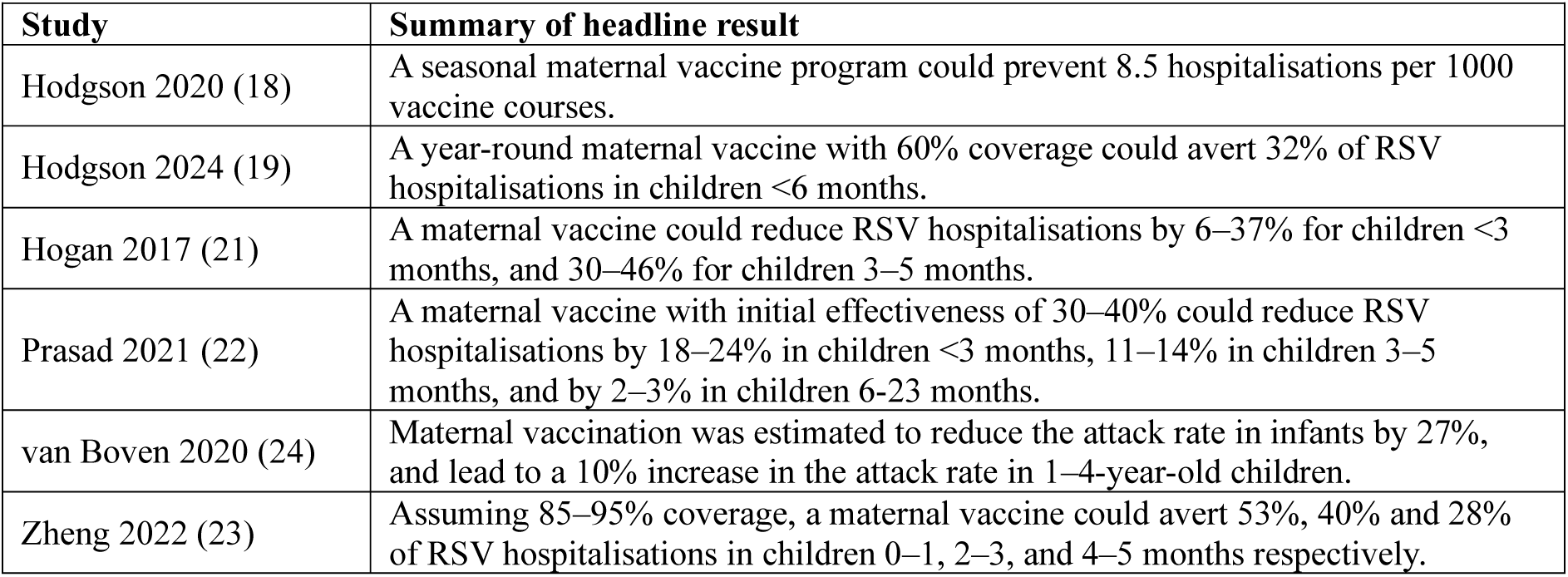
Summary of headline results for maternal vaccination studies.

**Table S2:** Summary of headline results for long-acting monoclonal antibody studies.

| Study | Summary of headline result |
| --- | --- |
| Hodgson 2020 (18) | A seasonal mAb program aimed at high-risk infants could prevent 36 hospitalisations per 1000 courses administered. |
| Hodgson 2024 (19) | A year-round mAb program with 90% coverage could avert 57% of RSV hospitalisations in children <6 months. |
| Prasad 2021 (22) | A seasonal infant mAb with 40–60% effectiveness for 150 days could reduce RSV hospitalisations by 30–43% in children <3 months, 34–48% in children 3–5 months, and by 14–21% in children 6–23 months. |
| Voirin 2022 (29) | Assuming 71% coverage and no impact on viral shedding, the average reduction in RSV-MALRTIs was 49.7% during the RSV season in infants <6 months, and 34.9% in infants aged 6–12 months. |
| Zheng 2022 (23) | Assuming 85–95% coverage, a mAb could avert 58%, 50% and 41% of RSV hospitalisations children 0–1, 2–3, and 4–5 months respectively. |

**Table S3:** Assumptions and parameters relating to implementation of natural maternally-derived immunity in the reviewed modelling studies. The study by van Boven et al did not explicitly capture maternally-derived protection in infants (24).

| Study | Type of modelled naturally-derived maternal immunity | Mechanism and parameterisation | References |
| --- | --- | --- | --- |
| Hodgson 2020 (18)<br>Hodgson 2024 (19) | Temporary complete immunity in a proportion of infants | Babies born to recently-infected mothers are born with temporary complete protection; average duration of 133.5 (119.6–146.1) days | Fitted; priors informed by (42–44) |
| Hogan 2017 (21) | Temporary partial immunity | Susceptibility to infection reduced by 92% in 1st month, and 55% in 2nd and 3rd months | Fixed; values informed by (45) |
| Prasad 2021 (22) | Temporary partial immunity | Susceptibility to infection reduced by 32% in first 3 months | Fitted; values informed by (45,46) |
| Voirin 2022 (29) | Temporary complete immunity | Average duration 2 months protection following birth | Fixed; values informed by (46) |
| Zheng 2022 (23) | Temporary complete immunity | Average duration $1/(\log N(-1,0.6))$ months protection following birth | Fitted; priors informed by (44,46–49) |

**Table S4:** RSV burden data to which reviewed models were calibrated.

| Study | Burden calibration data |
| --- | --- |
| Hodgson 2020 (18)<br>Hodgson 2024 (19) | Model fitted to age-stratified confirmed number of positive weekly RSV samples in England collected through respiratory data collection system, July 2010–June 2017. These were compared to the model-predicted output by multiplying modelled weekly incidence, with fitted age-specific reporting rates. |
| Hogan 2017 (21) | Model fitted to weekly age-stratified RSV laboratory-confirmed hospitalisations 2000–2017. Compared to model-predicted output by scaling modelled incidence by a fitted age-specific hospitalised proportion parameter. |
| Prasad 2021 (22) | Model fitted to weekly age-stratified RSV hospitalisations 2012–2015. Compared to model-predicted output by scaling modelled incidence by a fitted age-specific hospitalised proportion parameter. |
| van Boven 2020 (24) | Models fitted to 3 datasets: GP consultations for acute respiratory infections 2012–2017, weekly age stratified RSV hospitalisations 2013–2017, and test positivity fraction 2012–2016; compared to modelled RSV incidence and used to estimate age-stratified probabilities of GP consultation and hospitalisations. |
| Voirin 2022 (29) | Model fitted to incidence of age-stratified monthly RSV-MALRTI for years 2008–2015, with the reporting fraction estimated. |
| Zheng 2022 (23) | Model fitted to age-specific RSV hospitalisation data for 2005–2014, with the reporting fraction estimated from the fitting. |

## Notes

**Funding:** ABH is funded by an Australian National Health and Medical Research Council Investigator Grant (APP2009278). The contents of the published material are solely the responsibility of the authors and do not reflect the views of the NHMRC.

### Competing Interest Statement

ABH is a member of the World Health Organization Immunization and vaccines related implementation research advisory committee (IVIR-AC). Note that ABH an author on two of the studies included in the review.

### Funding Statement

ABH is funded by an Australian National Health and Medical Research Council Investigator Grant (APP2009278). The contents of the published material are solely the responsibility of the authors and do not reflect the views of the NHMRC.

